# Neural and Somatic Mechanisms Driving Clinical Improvements in Post-Acute Schizophrenia Spectrum Disorders

**DOI:** 10.1101/2024.09.27.24314427

**Authors:** Lukas Roell, Christoph Lindner, Isabel Maurus, Daniel Keeser, Berend Malchow, Andrea Schmitt, Peter Falkai

## Abstract

**Background:** A better mechanistic understanding of schizophrenia spectrum disorders is crucial to develop efficient treatment approaches. Therefore, this study investigated longitudinal interrelations between clinical outcomes, brain structure, and somatic health in post-acute individuals from the schizophrenia spectrum.

**Methods:** A sample of 63 post-acute patients from two independent physical exercise studies were included in the final analyses. Demographic, clinical, cognitive, and somatic data were acquired at baseline and post-intervention, as were structural magnetic resonance imaging scans. Multivariate cross-lagged panel modelling including mediators was used to study the mutual interrelations over time between the clinical, neural, and somatic level.

**Results:** A higher baseline global grey matter volume and larger regional grey matter volumes of the hippocampal formation, precuneus, and posterior cingulate drove improvements in multiple clinical outcomes, such as daily-life functioning, negative symptoms, and cognition. Increases in white matter volume from baseline to post-intervention resulted in significantly reduced positive symptoms and higher daily-life functioning following the intervention.

**Conclusion:** Our findings suggest that stimulating neuroplasticity, especially in the hippocampal formation, precuneus, and posterior cingulate gyrus, may represent a promising treatment target in post-acute schizophrenia spectrum disorders. Physical exercise therapies and other lifestyle interventions, and brain stimulation approaches reflect promising treatment candidates. Given the exploratory character of the statistical analysis performed, these findings need to be replicated in independent longitudinal imaging cohorts of patients with schizophrenia spectrum disorders.

## Introduction

Schizophrenia Spectrum Disorders (SSD) are debilitating psychiatric conditions characterized by positive, negative, and cognitive symptoms, as well as by long-term impairments in daily life functioning in a significant proportion of patients (1). Apart from those behavioral domains, patients with SSD are often affected by substantial somatic comorbidities, resulting in significant reductions in general body and organ health (2). Current treatment options encompass antipsychotic medication, psychotherapy, cognitive remediation, non-invasive brain stimulation, or physical exercise interventions, each differing with regard to their therapeutic windows and the underlying treatment goals (3–7). Despite the efficiency of these interventions, the current long-term disease outcomes in people with SSD offer substantial room for improvement: For instance, current estimates on remission rates averaged across different patient populations hover between 26 and 58 % (8,9), whereas recovery rates only range between 13.5 and 36 % (8–11). This places SSD among the top 20 diseases with the highest Years-Lived-With-Disability index (12), emphasizing the urgent need to improve current treatment options.

To enhance the long-term therapeutic success in SSD, a better mechanistic understanding of clinical symptoms, cognitive deficits, and daily-life functioning is warranted. Such mechanisms in SSD may be observable on the cerebral level assessed via structural magnetic resonance imaging (MRI). Large-scale cross-sectional evidence demonstrates that structural deteriorations in key brain regions such as the insula, anterior and posterior cingulate cortex, superior and middle frontal cortices, precuneus, hippocampal formation, putamen, pallidum, caudate, thalamus, and amygdala, or the cerebellum are linked to different domains of psychiatric symptoms and cognitive functioning in SSD (13–22). Moreover, widespread alterations in white matter integrity have been associated with cognitive deficits (23). However, these brain-behavior associations obtained from cross-sectional data yield only limited pathophysiological relevance, as they do not allow to make inferences on how the neural level affects the clinical level over time and vice versa.

To address this issue longitudinal neuroimaging studies are warranted. Respective evidence reveals an accelerated grey matter volume loss over time across multiple brain regions in SSD linked to the type of antipsychotic treatment and therefore potentially also to the long-term course of symptom severity (24–26). In line with these findings, grey matter volumes can serve as a predictor of long-term clinical outcomes in SSD (27), although many machine learning studies in this field have noticeable shortcomings such as a limited generalizability due to small sample sizes (28,29). In addition to methodological concerns, the exact mechanisms of action between brain structures and specific symptom domains in SSD remain largely unknown. For instance, in the context of physical exercise studies in SSD, there is preliminary evidence that exercise elicits diverse adaptations of brain structure throughout the brain, but the clinical implications remain unclear, as does the impact of clinical characteristics of patients on structural brain adaptability (30–34).

Besides the neural level, the significance of deteriorations in somatic health in SSD and other psychiatric conditions has been increasingly noticed in recent years (2). For instance, obesity in SSD is linked to both overall symptom severity (35) and impaired brain structure (36). However, the longitudinal interrelations between somatic health, clinical domains, and brain structure need to be further investigated in people with SSD.

Hence, we follow an exploratory research approach based on longitudinal data from two exercise intervention studies in SSD to first investigate the mutual interrelations between symptom severity, cognition, daily-life functioning, brain volumes, and somatic health at baseline and post-intervention. Second, we examine if changes in brain volumes and somatic health during the intervention mediate the temporal reciprocal interrelations between symptom severity, cognitive performance, and daily life functioning from pre- to post-intervention.

## Methods

This work utilizes data from two somatic exercise studies conducted in people with SSD. One is the Enhancing Schizophrenia Prevention and Recovery through Innovative Treatments (ESPRIT) C3 study (NCT03466112) performed at different sites across Germany (37,38). In the project at hand, only data acquired at the Department of Psychiatry and Psychotherapy of the Ludwig-Maximilians-University Hospital in Munich was utilized. The second exercise trial (NCT01776112) was executed at the Department of Psychiatry and Psychotherapy of the University Medical Center Goettingen (30,39).

Both studies were in line with the Declaration of Helsinki and ethical approval was provided by the local ethics committees of the Ludwig-Maximilians-University Hospital and the University Medical Center Goettingen, respectively.

### Sample and Study Design

The ESPRIT C3 study investigated the effects of two different types of exercise on several health outcomes in people with SSD. Patients were either randomized to an aerobic exercise intervention on bicycle ergometers or to a flexibility, strengthening, and balance training. Both groups exercised three times per week between 40 and 50 minutes over a period of six months. For this work, we considered behavioral and MRI data from pre- and post-intervention. Details on the study design and the main results are described elsewhere (37,38)

The second study entailed a three-month aerobic exercise intervention on bicycle ergometers with additional cognitive remediation starting in the sixth week of the intervention. Patients exercised three times per week for 30 minutes per session. The control group played table soccer for the same amount of time and also received cognitive remediation after six weeks of the intervention. The project at hand considers behavioral and MRI data from baseline and post-intervention only of the patients diagnosed with SSD.

Inclusion and exclusion criteria are listed in the original publications (30,37–39). In the ESPRIT C3 study only a subgroup of patients underwent the MRI assessments. After quality control of behavioral and MRI data (for details see supplemental information), 63 people with SSD were included in the final analysis.

### MRI Data Acquisition

Participants of the ESPRIT C3 study were scanned in a 3T Siemens Magnetom Skyra MRI scanner (SIEMENS Healthineers AG, Erlangen, Germany) at the Department of Radiology of the Ludwig-Maximilians-University Hospital Munich. A 3D T1-weighted magnetization-prepared rapid gradient echo (MPRAGE) sequence with an isotropic spatial resolution of 0.8 x 0.8 x 0.8 mm^3^ was acquired. In the second exercise study, a 3T Magnetom TIM Trio MRI scanner (SIEMENS Healthineers AG, Erlangen, Germany) was used to acquire a 3D T1-weighted MPRAGE sequence at 1.0 x 1.0 x 1.0 mm^3^ isotropic resolution. Supplementary Table S1 summarizes the scanning parameters.

### Quality Control and MRI Data Processing

All available structural images were inspected visually to evaluate the data quality. The automated quality control software MRIQC was used compute relevant image quality metrics (40). Further details on the quality control procedure are provided in the supplemental information.

Structural images were processed using the Neuromodulation and Multimodal Neuroimaging Software (NAMNIs) version 0.3 (41). NAMNIs is mainly based on tools from FSL (42) and includes the following processing steps for structural MRI data: image reorientation to standard space, brain extraction, creation of a binary masks, linear and non-linear registration, inversion of the transformation and deformation field, brain segmentation, computation of global grey matter volume, white matter volume and cerebrospinal fluid, mapping of brain atlas of choice into the native space, and calculation of regional grey and white matter volumes.

We used the third version of the Automated Anatomical Labeling (AAL3) atlas (43) to compute the regional grey matter volumes of the bilateral insula, anterior and posterior cingulate cortices, superior and middle frontal cortices, precuneus, hippocampus, putamen, pallidum, caudate, thalamus, and amygdala. The aformentioned regions were selected based on previous literature suggesting their clinical relevance in SSD (see introduction). The regions of interest are visualized in Figure 1.

**Figure 1:**
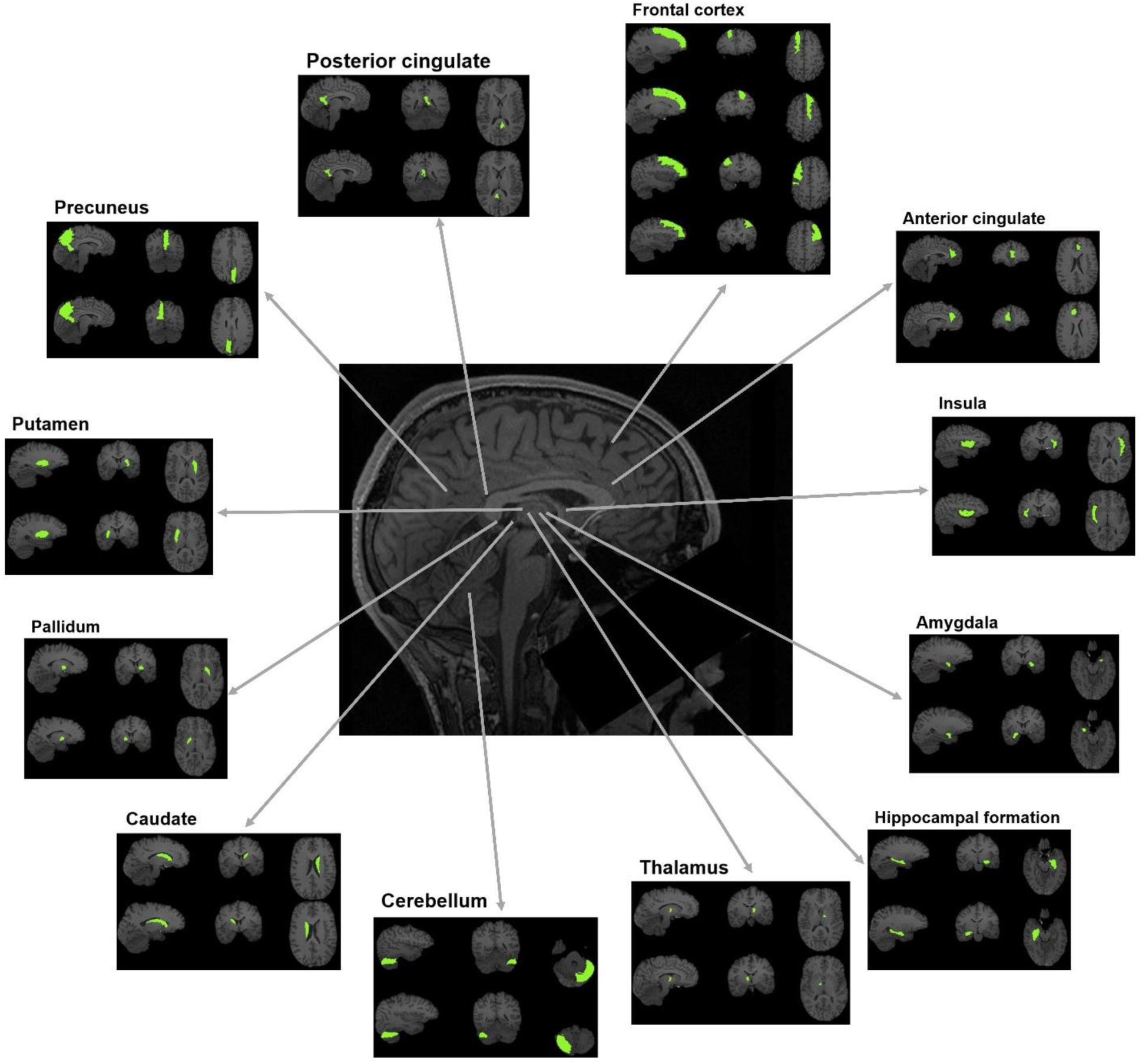
Regions of Interest. The regions of interest are visualized for which grey matter volumes were computed. Note that for the thalamus, cerebellum, and anterior cingulate gyrus only one subregion is illustrated in this figure. For the analyses, the volumes of these subregions were summed up to one volume score.

### Clinical and Cognitive Assessments

To assess positive and negative symptoms the Positive and Negative Syndrome Scale (PANSS) (44) was utilized in both exercise studies, as was the Global Assessment of Functioning (GAF) scale to assess daily life functioning (45). The first trial and the interference trial of the Verbal Learning and Memory Test (VLMT) (46) were averaged to a short-term memory score, whereas the sixth and seventh trial were averaged to a long-term memory score. The backward version of the Digit Span Test (DST) (47) was used to assess working memory. The Trail Making Tests (TMT) A and B (48) were averaged to provide a more global measure of cognition covering multiple domains such as processing speed, working memory updating, and response inhibition. A detailed description of the cognitive test batteries is provided in the supplemental information. Clinical assessments and cognitive tests were administered pre- and post-intervention in both exercise studies.

### Somatic Health Assessments

To quantify general somatic health, we computed a principal component analysis across subjects and sessions including body-mass-index (BMI), and levels of cholesterol, HbA1c, and triglycerides as variables. We used the score on the first principal component of each subject in each session as an indicator of general health. A lower score on the first principal component was associated with a higher BMI and higher levels of cholesterol, HbA1c, and triglycerides, thus indicating a worse somatic health (supplemental information).

### Statistical Data Analysis

To achieve the first aim of the study, namely studying the reciprocal temporal relations between symptom severity, cognition, daily life functioning, brain volumes, and somatic health measured at baseline (first measurement occasion) and after the intervention (second measurement occasion), multivariate cross-lagged panel modelling (CLPM) (49) was applied using the lavaan package (50) in R version 4.2.2. We fitted a total of 28 separate models for global grey and white matter volume, as well as for 13 brain regions of interest for both hemispheres. In all our models, (residual) correlations between variables at the same levels (i.e. pre- and post-intervention) were allowed. Further, the models comprised exclusively manifest variables as indicators, represented in positive and negative symptom severity scores, short-term, long-term, and working memory performance scores, daily life functioning scores, somatic health scores, and the respective brain volume scores, each at baseline and post-intervention. In all CLPM, the autoregressive paths represented the stability of individual differences from the first to the second measurement occasion. These are the effects of all our measured variables on themselves. The cross-lagged paths are the effects of each variable on each other variable measured at a later timepoint, while controlling for the prior level of the corresponding variable being predicted (51). Thus, the CLPM allowed us to explore the directional interrelations between all variables of investigation over the course of time. Given the exploratory approach of this study, no correction for multiple comparisons was performed, but significant results of single brain regions were only further interpreted if they were consistent across hemispheres. Figure 2A visualizes the general structure of the cross-lagged panel models.

**Figure 2:**
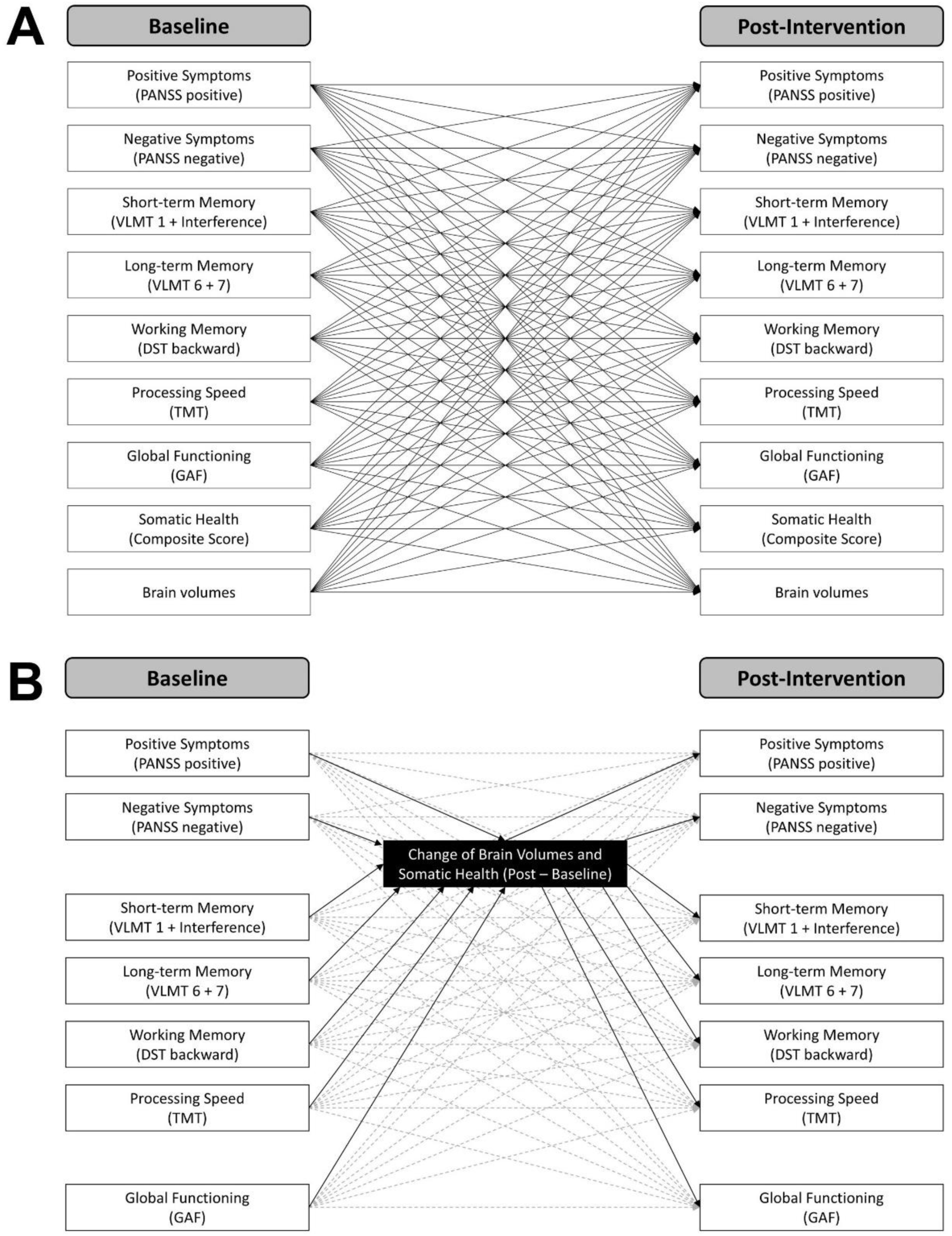
Cross-lagged Panel Models. The manifest path models computed in the current study are illustrated. A) Cross-lagged panel models including baseline and post-intervention scores. The global and regional brain volumes were inserted separately. Note that covariances between baseline and between post-intervention variables and covariates are not displayed for the sake of visibility. B) Cross-lagged panel models with mediators including clinical and cognitive variables at baseline and post-intervention, as well as brain volumes and somatic health as change scores (post-intervention – baseline). Autoregressive and cross-lagged paths are illustrated in dashed grey lines. Note that covariances between baseline and between post-intervention scores are not shown for the sake of visibility.

Following the second aim of the study, examining the potential mediating role of brain volume and somatic health changes during the intervention, we included additional manifest mediator variables in our CLPM. More precisely, we specified the CLPM as described above and additionally included the change of brain volumes and somatic health scores (i.e., post-intervention - pre-intervention) as mediators. We defined a total of 29 separate models for the change in somatic health score, global grey and white matter volume, as well as for changes in 13 brain regions of interest. Beside the two mediator variables, the models contained positive and negative symptom severity scores, short-term, long-term, and working memory performance scores, and daily life functioning scores at baseline and post-intervention. Specifying CLPM with additional mediator variables allowed us to investigate the role of changes in brain volume somatic health in the relation between baseline and post-intervention levels of the investigated variables while simultaneously controlling for any additional effects of the corresponding variables in the models. No correction for multiple comparisons was performed. Significant results in the case of single brain regions were only considered if they were consistent across hemispheres. Figure 2B illustrates the general structure of the mediation models.

In all our models, age, sex (female = 0, male = 1), chlorpromazine equivalents (Defined Daily Doses method (52)), years of education, exercise group (first dummy coding: aerobic exercise = 0, flexibility, strengthening, and balance training = 1; second dummy coding: aerobic exercise = 0, table soccer = 1), and study (Goettingen = 0, Munich = 1) were included as covariates to control for confounding influences of these variables on the interrelations between the main variables of investigation. Furthermore, if not otherwise described the relations between all variables and covariates were allowed to vary freely in all models. Thus, all our models were saturated with *df* = 0, meaning that the models had as many freely estimated parameters as observations in the data set (i.e., variances, covariances, means). The maximum likelihood estimator was used and confidence intervals were calculated. The analysis dataset was free of missing data.

## Results

### Patients Characteristics

Table 1 summarizes the sample characteristics. The majority of subjects received an aerobic exercise therapy. More males than females were included. Overall, the sample was at a rather post-acute phase of the disease, as indicated by relatively low positive symptom severity and average functioning scores.

**Table 1.** Sample Characteristics.

| | **Patients with SSD**<br>**N = 63**<br>(n/mean ± SD) |
| --- | --- |
| **Study** |  |
| Goettingen | 42 |
| Munich | 21 |
| **Study arm** |  |
| Aerobic exercise | 27 |
| Flexibility, strengthening, and balance training | 15 |
| Table soccer | 21 |
| **Sex** |  |
| Female | 19 |
| Male | 44 |
| **PANSS at baseline** |  |
| Positive symptoms | 12.50 ± 5.60 |
| Negative symptoms | 16.30 ± 8.20 |
| **GAF at baseline** | 61.9 ± 11.0 |
| **Chlorpromazine equivalents** | 526 ± 524 |
| **Education (years)** | 15.4 ± 3.92 |
| **Total number of trainings** | 33.7 ± 9.20 |

The sample size refers to the number of participants that were considered in the final statistical data analysis. N, total sample size; n, sample size per category; PANSS, Positive and Negative Syndrome Scale; GAF, Global Assessment of Functioning Scale; SD, standard deviation.

### Reciprocal Longitudinal Interrelations between Clinical, Neural, and Somatic Outcomes

Across all 28 cross-lagged panel models, multiple significant paths were detected: All autoregressive paths were significant (Table S3). With regard to the cross-lagged paths, a higher global grey matter volume predicted higher working memory performance post-intervention (β = 0.36, CI = [0.09, 0.63], p = 0.010), whereas global white matter volume had no effect. With regard to the single brain regions, a higher bilateral hippocampal grey matter volume at baseline drove higher post-intervention daily life functioning (left: β = 0.21, CI = [0.04, 0.39], p = 0.018; right: β = 0.19, CI = [0.01, 0.38], p = 0.042). Larger grey matter volume in the bilateral precuneus resulted in lower negative symptom severity (left: β = −0.26, CI = [-0.09, −0.42], p = 0.002; right: β = −0.19, CI = [-0.02, −0.36], p = 0.027), higher working memory performance (left: β = 0.28, CI = [0.04, 0.51], p = 0.019; right: β = 0.35, CI = [0.13, 0.58], p = 0.002), and higher somatic health (left: β = 0.22, CI = [0.04, 0.40], p = 0.017; right: β = 0.23, CI = [0.05, 0.41], p = 0.011) after the intervention. Larger grey matter volume in the posterior cingulate gyrus predicted higher post-intervention long-term memory performance (left: β = 0.18, CI = [0.03, 0.33], p = 0.021; right: β = 0.20, CI = [0.06, 0.34], p = 0.004) and higher post-intervention levels of daily-life functioning (left: β = 0.30, CI = [0.11, 0.48], p = 0.001; right: β = 0.20, CI = [0.02, 0.37], p = 0.031). A higher baseline grey matter volume in the bilateral insula resulted in more severe positive symptoms at post-intervention (left: β = 0.22, CI = [0.04, 0.40], p = 0.017; right: β = 0.20, CI = [0.04, 0.37], p = 0.017). A higher somatic health at baseline drove larger grey matter volume in the bilateral cerebellum (left: β = 0.09, CI = [0.01, 0.18], p = 0.038; right: β = 0.15, CI = [0.03, 0.26], p = 0.013), while a higher working memory performance (left: β = 0.10, CI = [0.02, 0.17], p = 0.015; right: β = 0.09, CI = [0.01, 0.17], p = 0.024) and daily-life functioning (left: β = 0.14, CI = [0.02, 0.26], p = 0.021; right: β = 0.14, CI = [0.02, 0.26], p = 0.028) at baseline resulted in larger bilateral grey matter volume in the caudate nucleus. Similarly, a higher working memory performance at baseline predicted larger grey matter volume in the bilateral putamen (left: β = 0.13, CI = [0.01, 0.24], p = 0.029; right: β = 0.14, CI = [0.00, 0.27], p = 0.045). No other brain regions revealed similar effects across both hemispheres. A higher positive symptom severity at baseline resulted in worse post-intervention somatic health across all 28 models (for model with global grey matter volume as neural entity: β = −0.25, CI = [- 0.06, −0.43], p = 0.010). In some models a higher somatic health at baseline was linked to a higher working memory performance after the intervention (supplemental table S2), but given the inconsistency of this finding we did not further interpret it. Supplemental table S2 and S3 summarize all statistics including overall model fits and effects sizes, confidence intervals and p-values for all significant and non-significant paths.

### Mediating Role of Changes in Brain Volumes and Somatic Health

Across all 29 cross-lagged panel models with mediators, only a few significant paths were obtained: All autoregressive paths were significant (Table S5 and S7). An increase in global white matter volume from pre- to post-intervention drove lower positive symptom severity (β = −0.64, CI = [−0.05, −1.23], p = 0.033) and higher daily-life functioning (β = 0.77, CI = [0.02, 1.51], p = 0.044) after the intervention, while a worsening in somatic health was linked to higher daily life functioning (β = −0.34, CI = [-0.10, −0.57], p = 0.005) following the intervention. A higher working memory performance at baseline resulted in a more pronounced increase of grey matter volume in the bilateral caudate nucleus (left: β = 0.09, CI = [0.01, 0.17], p = 0.022; right: β = 0.09, CI = [0.00, 0.17], p = 0.043). No further brain regions showed consistent mediating effects across both hemispheres. Supplemental tables S4 – S7 contain all statistics including overall model fits and effects sizes, confidence intervals and p-values for all significant and non-significant paths.

## Discussion

Inspired by the long-term aim to identify potential treatment targets in SSD, the current study first explored the reciprocal interrelations between symptom severity, cognition, daily-life functioning, brain volumes, and somatic health before and after the intervention. Further we investigated if changes in brain volumes and somatic health mediate the mutual interrelations of clinical symptoms, cognitive performance, and daily-life functioning from pre- to post-intervention.

We found that a higher global grey matter volume at baseline resulted in higher working memory performance following the intervention. This is in line with current findings in the general population, demonstrating that higher initial grey matter volume mitigates age-related cognitive decline over time, while long-term grey matter volume reductions throughout the lifespan are directly associated with continuous cognitive decline (53). In people with SSD, global grey matter volume is reduced compared to controls (54) and an accelerated loss of grey matter over time has also been demonstrated (24–26). Considering the concept of cognitive reserve (55), these findings point toward a decreased brain reserve in SSD potentially characterized by reduced numbers of neurons or synapses throughout the brain, increasing the proneness to cognitive impairment. Our findings fit well to this framework, as we could show that those patients with higher global grey matter volume will show higher working memory performance after the intervention. Interestingly, we did not observe similar effects for other cognitive domains which may reflect an issue of statistical power, because when using a global cognition score as clinical outcome, we obtained the effect again (supplemental table S9). In sum, our results highlight the importance of preventing global grey matter volume decline in SSD, because patients with pronounced grey matter loss are less likely to show cognitive benefits in the context of an intervention. Therefore, future treatments in SSD should target general brain health, while also considering somatic health, given the influence of the latter on brain structure in SSD (36). Combined lifestyle interventions targeting physical activity, diet, sleep, and substance use may be promising candidates to tackle brain and somatic health in SSD, as indicated by respective evidence published in recent years (56–61).

Our results further suggest that a larger hippocampal grey matter volume at baseline predicts higher daily-life functioning following the intervention. Structural impairments of the hippocampal formation in SSD has been repeatedly demonstrated (14,22,54,62–67). With regard to the clinical implications of hippocampal volume decline in SSD, converging evidence reveals associations to impaired cognitive functioning (18,68), to symptomatic worsening over time (69), and to long-term deteriorations in daily-life functioning (70). The current results build on these findings, indicating that patients with higher baseline grey matter volume in the hippocampal formation show higher daily-life functioning after the intervention. This effect may be driven by the following mechanisms: Decreased hippocampal volume in SSD observed in neuroimaging studies is sought to result from atrophy processes involving a reduction of the number, volume and size of neurons (71) and oligodendrocytes (72,73). This atrophy may origin from elevated glutamate levels within the hippocampal formation in SSD that cause a spreading pattern of hypermetabolism and pronounced excitation-inhibition imbalance (74). Imbalanced excitatory and inhibitory signaling throughout the brain represents a core dysregulation, contributing to the general pathology in SSD (75). Hence, excitation-inhibition imbalance in the hippocampal formation may underly long-term reductions of grey matter volume and thus explain the effect on daily-life functioning observed in our data. To conclude, our findings emphasize the importance of hippocampal health in SSD impacting the capability of patients to show improvements in daily-life functioning. Consequently, future treatments should aim for stimulating hippocampal neuroplasticity to ameliorate the global functional outcome of patients. Aerobic exercise interventions (56) or – possibly at some future stage – subcortical brain stimulation methods such as focused ultrasound (76) may reflect a promising candidate for such treatment approach.

Our findings further indicate that a larger grey matter volume in the precuneus at baseline explains lower negative symptoms, higher working memory performance, and higher somatic health after the intervention, while a higher grey matter volume in the posterior cingulate gyrus leads to higher long-term memory performance and daily-life functioning. Both the precuneus and the posterior cingulate gyrus, but also the previously discussed hippocampal formation, are part of the default-mode network reflecting a self-referential introspective neural state linked to theory of mind and social cognition (77,78). In SSD, both structural and functional abnormalities in the default-mode network align with more severe negative symptom severity (79,80), impairment in theory of mind (81) and decline in working memory (82). Furthermore, grey matter volume of the posterior cingulate gyrus has been shown to predict functional outcome in SSD (83), while functional segregation and integration within the default-mode network has been suggested to influence treatment response (84). Our findings align with these results, emphasizing the crucial role of the default-mode network in the pathophysiology of SSD. Future studies should evaluate possibilities to influence structure and function of the default-mode network to improve the overall psychiatric health status of post-acute patients with SSD. Different types of neurostimulation methods may be promising candidates (76,85).

Our findings suggest that increases in global white matter volume from pre- to post-intervention drive lower positive symptom severity and higher daily-life functioning after the intervention. White matter pathology in SSD has been mostly associated with cognitive deficits, both on the empirical (23) and the theoretical level (86). Our data rather point towards its relevance for positive symptom severity and global functioning, but especially the latter strongly depends on cognitive functioning (87). Hence, increasing global white matter volume and improving myelination of the brain in SSD can be regarded as a promising treatment approach to achieve further global improvements in symptomatology, cognition, and daily-life functioning, Indeed, a recent theory suggests a deficient maturation of oligodendrocyte precursor cells to cause cognitive deficits in SSD and proposes a combined therapy of aerobic exercise and clemastine to improve myelin plasticity and global structural connectivity (86). Further results of this study are discussed in the supplemental information.

Our study includes several limitations that have important implications for future research. First, we did not correct for multiple comparisons, as we aimed to conduct a global exploratory analysis to generate hypotheses on regarding the mutual interrelations between clinical outcomes, brain volumes, and somatic health. Hence, our findings require a replication in an independent sample to evaluate if the identified mechanisms of action are stable across samples. Future longitudinal imaging studies in SSD should use hypothesis-driven approaches, focusing for instance on the relevant regions identified in the current work. Second, the directional effects found in this study need to be interpreted with caution. Even though we used cross-lagged panel modeling to study reciprocal effects, there may have been confounding variables unavailable in the current datasets, such as medication change, that may explain the effects obtained. Future studies need to repeat the current analyses using a randomized-controlled design while drawing on a larger sample to enable robust causal inferences. Lastly, our findings have limited generalizability, as the underlying sample mainly consist of stable and post-acute patients with SSD who participated in different exercise intervention studies. Consequently, the observed mechanisms may only be present in post-acute SSD and may be specific in the context of exercise treatments. Hence, replication is required in other patient populations that were assessed in different longitudinal study designs to evaluate if the current findings reflect general pathophysiological trajectories.

To conclude, our results suggest that increasing the global grey and white matter volume and the regional grey matter volumes in the hippocampal formation, precuneus, and posterior cingulate gyrus reflect potential treatment targets to achieve further clinical improvements in post-acute SSD. Multiple treatment approaches reflect promising candidates to address these targets, although a replication of the present findings in independent cohorts is warranted before administering respective interventions in patient populations.

## Data Availability

All data produced in the present study are available upon reasonable request to the authors.

## Acknowledgments

The work was supported by the German Federal Ministry of Education and Research (BMBF) through the research network on psychiatric diseases ESPRIT (Enhancing Schizophrenia Prevention and Recovery through Innovative Treatments; coordinator: Andreas Meyer-Lindenberg; grant number, 01EE1407E) to PF and AS. Furthermore, the study was supported by the Else Kröner-Fresenius Foundation with the Research College “Translational Psychiatry” to PF, AS, and IM (Residency/PhD track of the International Max Planck Research School for Translational Psychiatry [IMPRS-TP]). The study was endorsed by the Federal Ministry of Education and Research (Bundesministerium für Bildung und Forschung [BMBF]) within the initial phase of the German Center for Mental Health (DZPG) (grant: 01EE2303A, 01EE2303F to PF, AS).

The authors express their appreciation to the Clinical Open Research Engine (CORE) at the University Hospital LMU (Munich, Germany) for providing the computational infrastructure to run the CPU-intensive MRI analysis pipelines.

