## Supplemental Information for "Neural and Somatic Mechanisms Driving Clinical Improvements in Post-Acute Schizophrenia Spectrum Disorders"

### Supplementary Information

#### Quality Control of Behavioral and Imaging Data

All demographic, clinical, cognitive, and physical variables were evaluated with regard to implausible values resulting from erroneous data entry or other factors. Figures S1 – S3 visualizes the data distributions of all relevant variables.

Structural imaging data were inspected visually and rated with regard to their quality. Figure S4 displays three examples of T1w images of different quality. Additionally, images were further quality controlled using the automated software MRIQC (1). The distributions of the image quality metrics were plotted and subjects with outlying metrics in a specific session were further inspected visually. In sum, six subjects were excluded due to lacking data quality resulting in the final sample size of 63 subjects. After excluding these subjects, the distributions of gray matter volumes were visualized (Figure S5) to check for severe outliers. Those very few single values were imputed using k-nearest neighbors (see left pallidum as an example in Figure S5).

**Figure S1**: Demographic variables

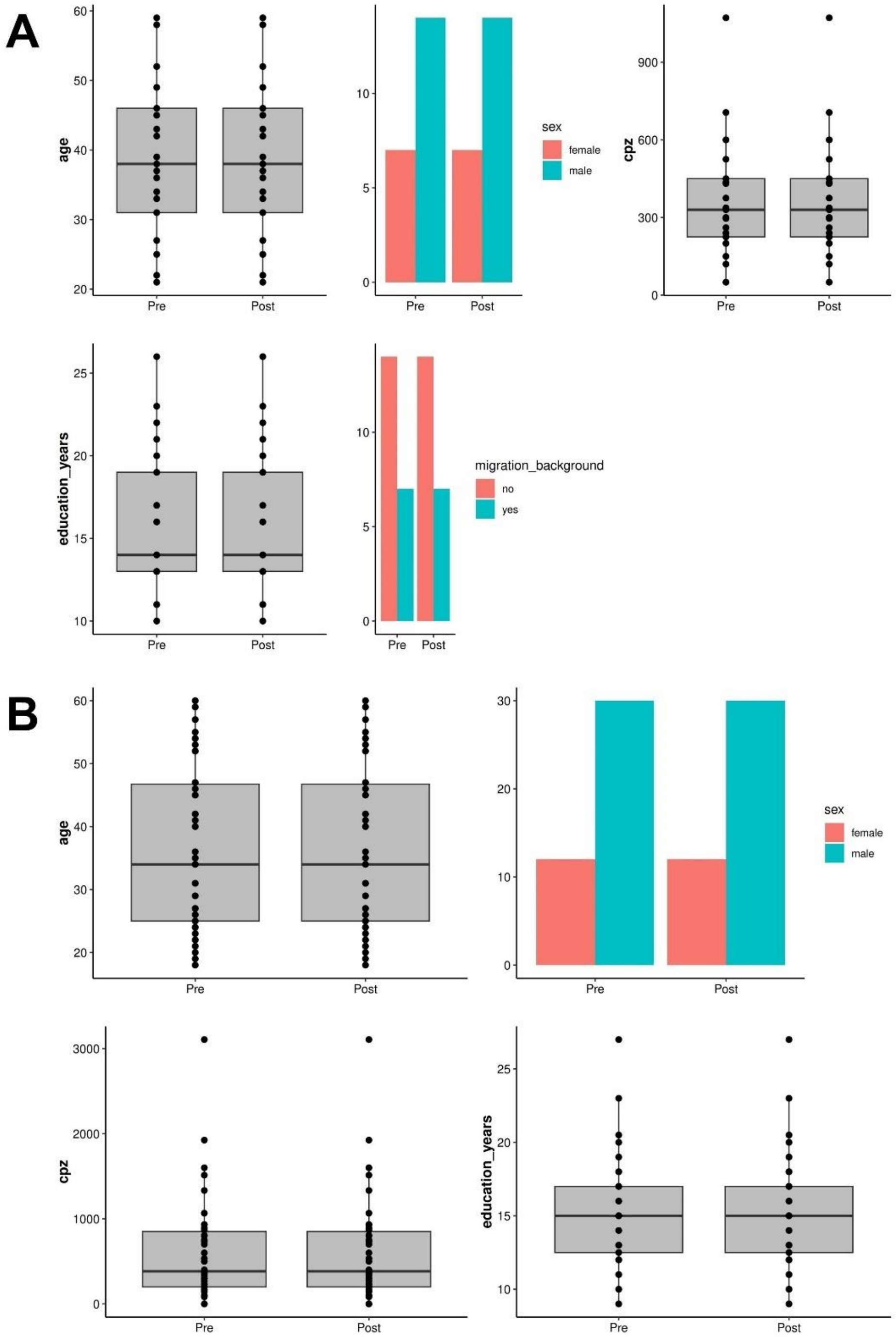


Data distributions of all demographic variables that were of interest in the current study. A) Data of ESPRIT C3 Exercise study conducted in Munich. B) Data of Exercise 2 study conducted in Goettingen. Note that migration background was only available in the ESPRIT study and therefore was not considered in the structural equation models.

**Figure S2**: Clinical and cognitive variables

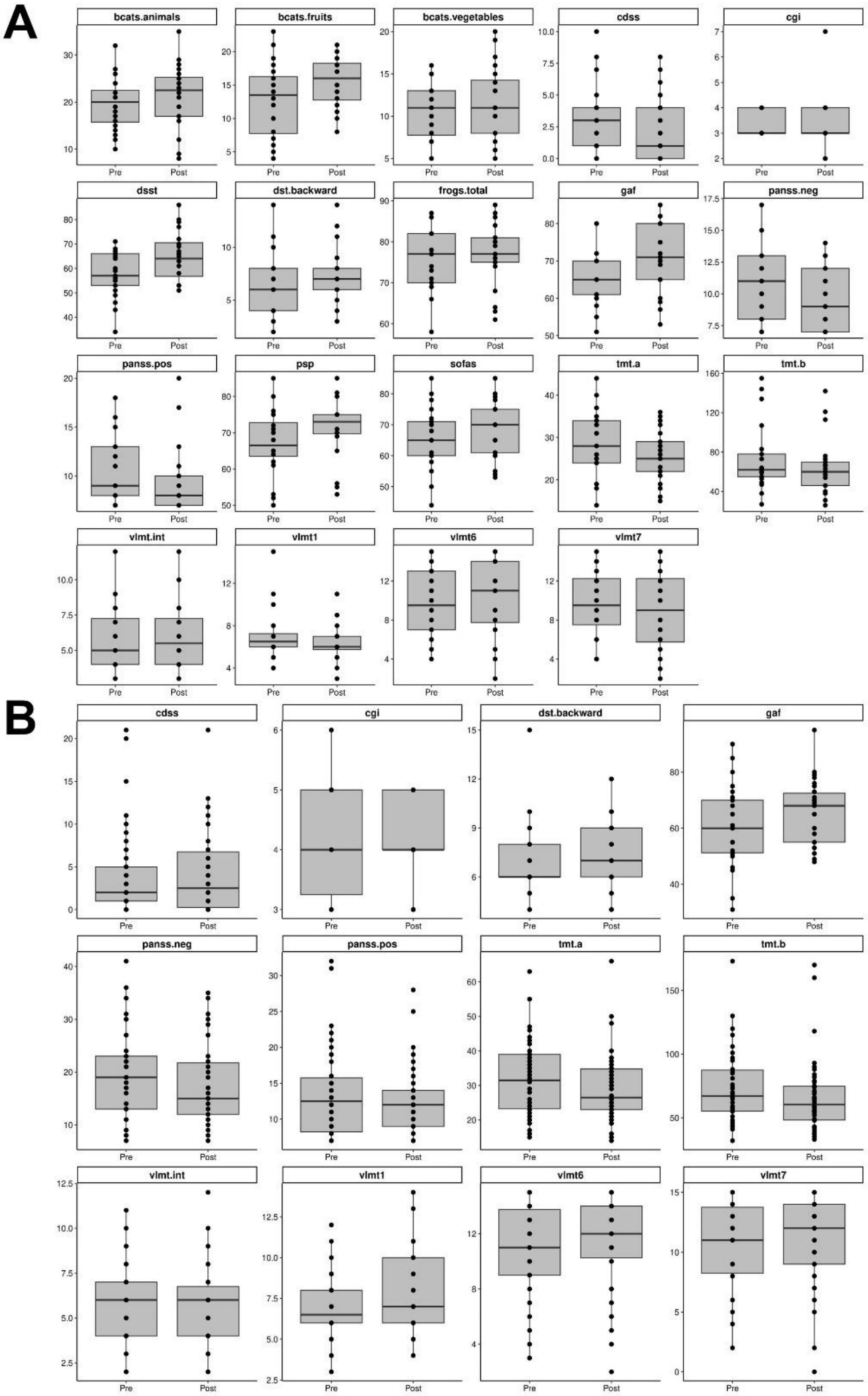


Data distributions of all clinical and cognitive variables that were of interest in the current study. A) Data of ESPRIT C3 Exercise study conducted in Munich. B) Data of Exercise 2 study conducted in Goettingen. Note that several variables were only available in the ESPRIT study and therefore were not considered in the structural equation models.

**Figure S3**: Physical health variables

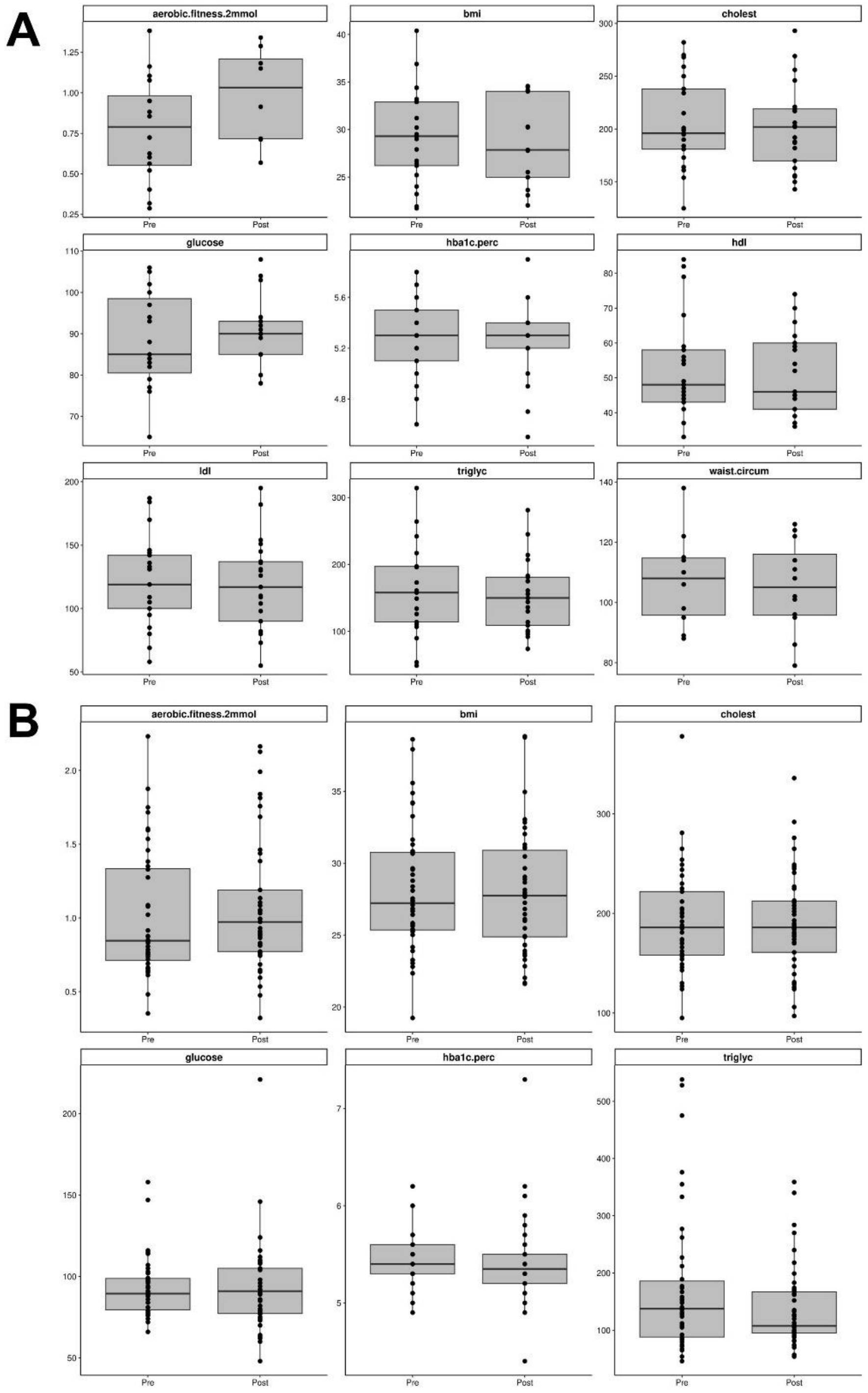


Data distributions of all physical health variables that were of interest in the current study. A) Data of ESPRIT C3 Exercise study conducted in Munich. B) Data of Exercise 2 study conducted in Goettingen. Note that several variables were only available in the ESPRIT study and therefore were not considered in the structural equation models.

**Figure S4**: Examples of good and bad image quality

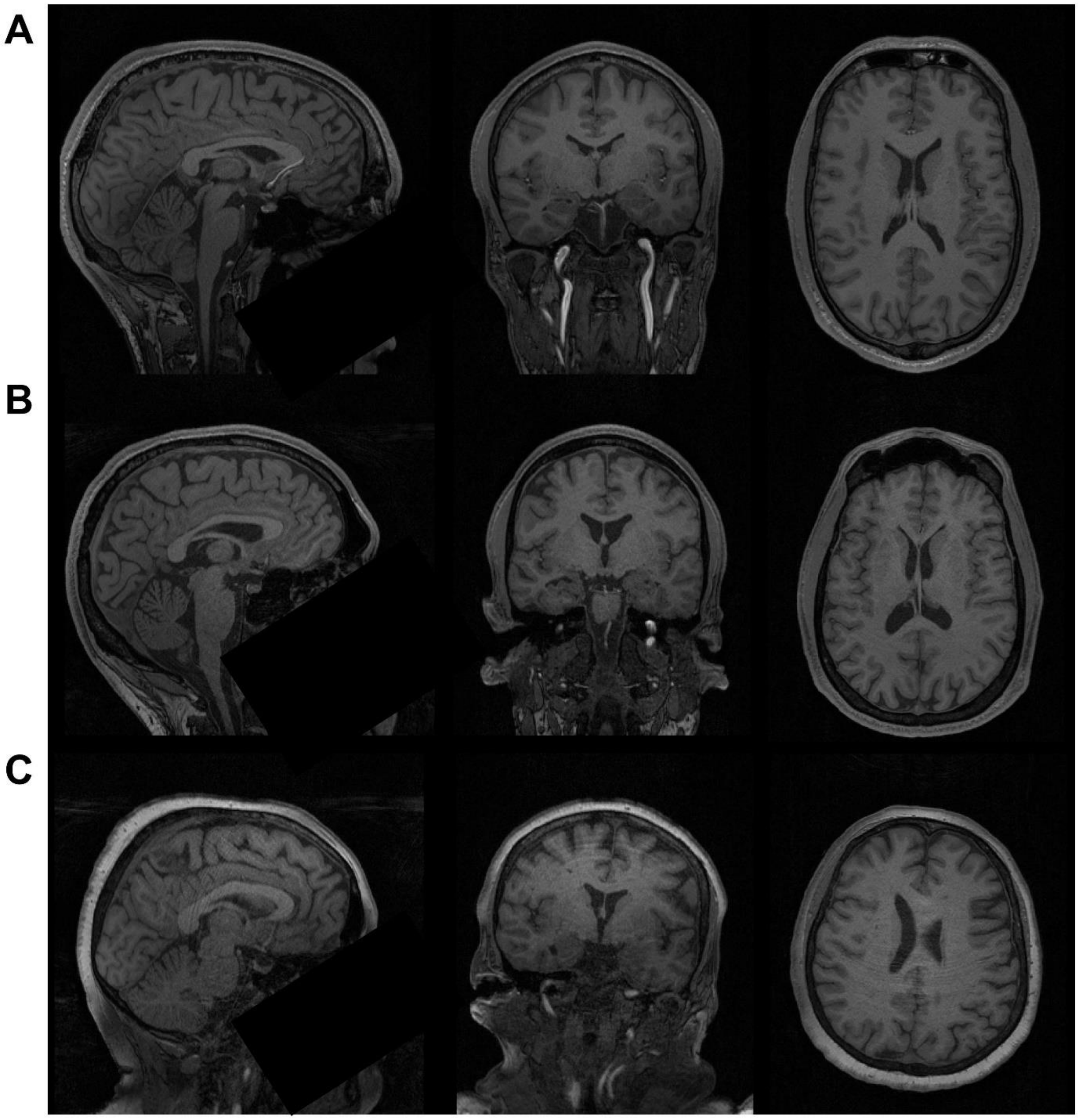


Three examples of T1w images with different data quality. A) Acceptable image that would be considered for further analyses. B) Image with medium quality given the visible motion artifacts. Such images were excluded. C) Image with bad quality given the severe motion artifacts. Such images were excluded.

**Figure S5**: Brain volumes

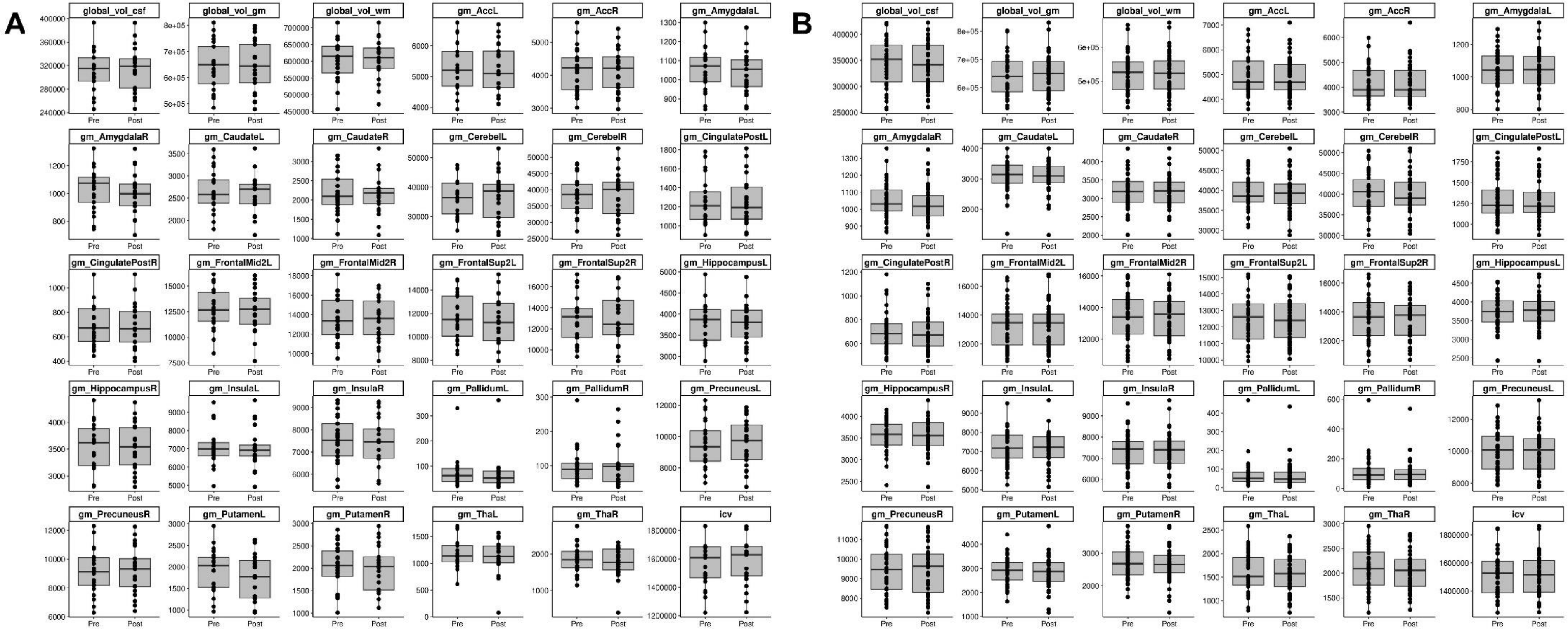


Data distributions of all brain volumes that were of interest in the current study. A) Data of ESPRIT C3 Exercise study conducted in Munich. B) Data of Exercise 2 study conducted in Goettingen.

**Scanning parameters**

**Table S1**. Scanning parameters.

| Site | Sequence | Resolution | TR | TE | TI | FA | Slices |
|---|---|---|---|---|---|---|---|
| Munich | MPRAGE | $0.8 \times 0.8 \times 0.8$ mm³ | 2060 ms | 2.20 ms | 1040 ms | 12° | 256 |
| Goettingen | MPRAGE | $1 \times 1 \times 1$ mm³ | 2250 ms | 3.26 ms | 900 ms | 9° | 176 |

Site, study site; Sequence, type of scanning sequence; FoV, field of view; resolution, voxel size; TR, Time of repetition; TE, echo time; TI, inversion time; FA, flip angle; slices, number of acquired slices; MP-RAGE, T1-weighted magnetization prepared rapid gradient echo; EPI, echo planar imaging.

#### Cognitive Test Batteries

We administered the Verbal Learning and Memory Test (VLMT) (2), Digit Span Test (DST) (3), and the Trail Making Test (TMT) A and B (4)

Within the VLMT the investigator reads a list of 15 words and the participants had to remember as many words as possible in arbitrary order. This procedure was repeated five times in a row (VLMT-1$^{st}$ to VLMT-5$^{th}$) and the sum of correctly remembered words across the five trials was computed (VLMT-sum). After the fifth trial, an interference list of 15 different words was read and the subjects had to name as many words from this new list as possible (VLMT-inter). The subject was asked to remember as many words as possible from the first list (VLMT-6$^{th}$) without repeating it again. After a 20 minutes delay in which other cognitive tests were executed, the participants had to remember as many words as possible from the first list again (VLMT-7$^{th}$). The difference in remembered words between VLMT-5$^{th}$ and VLMT-7$^{th}$ was calculated and multiplied by minus one (VLMT-diff). Finally, the investigator reads 50 words including the ones from the first list and the interference trial and the subjects had to decide if the corresponding word was part of the first list (VLMT-recog).

The number of correctly remembered or recognized words in each trial was counted and z-standardized resulting in eight different VLMT-scores.

During the DST-forward the investigator read digit rows of increasing lengths that the subject had to repeat verbally in the same order. The test was stopped if the participant failed twice within the same “row length category”. DST-backward worked analogously, except that the subjects had to repeat the digits in reverse order. The number of correct trials was counted, z-standardized in both versions separately

During the TMT-A subjects had to connect numbers from 1 to 25 in ascending order as fast and accurately as possible without lifting the pencil from the sheet of paper. In TMT-B participants had to connect numbers and letters alternately in the following order until number 13 was reached: 1-A-2-B-3-C-4-D-5-E-6-F-7-G-8-H-9-I-10-J-11-K-12-L-13. The time needed in seconds was measured. Results from both versions were multiplied by minus one, z-standardized across sessions and participants and averaged to a global TMT score.

#### Physical Health Score

Figure S6 reveals the correlation between the global physical health score on the one hand and the BMI, cholesterol, HbA1c, and triglycerides on the other hand, illustrating that a higher physical health score was reflecting a better general physical health. All correlations were highly significant with $p < 0.001$.

**Figure S6**: Correlations between physical health score and original variables

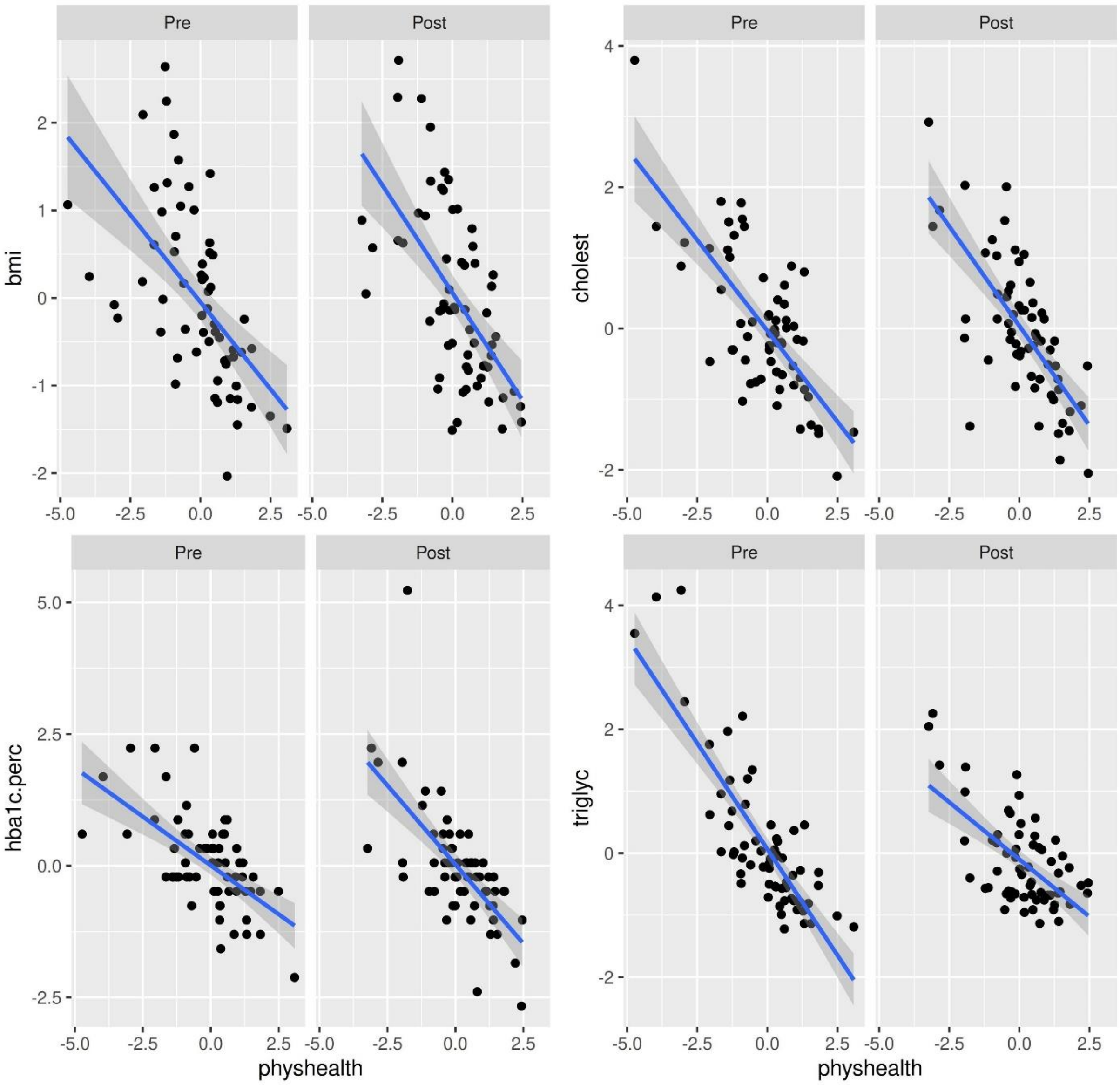


Correlations between physical health score on the x-axis and the respective original variable on the y-axis (BMI, cholesterol, HbA1c and triglycerides) that contributed to the physical health score. Pre and Post refer to pre- and post-intervention.

#### Test Statistics

Tables S2 – S7 summarize the whole test statistics for the cross-lagged panel and change models. The tables are provided as Excel files.

#### Alternative Cross-lagged Panel Model including Global Cognition

Tables S8 and S9 show the test statistics of an alternative cross-lagged panel model investigating the impact of global gray matter volume at baseline on global cognition after the intervention. The tables are provided as Excel file.

**Supplemental Discussion**

In addition to the findings discussed in the main manuscript, we further observed that better somatic health at baseline predicted larger grey matter volume in the cerebellum after the intervention. To our knowledge, this is the first time such finding has been reported. Given the role of the cerebellum in motor functioning (5), this result may suggest that patients in a better somatic condition are able to engage more efficiently in a physical exercise intervention leading to enhanced neuroplasticity in cerebellar motor systems.

In addition, we obtained that the patients with better baseline working memory performance and daily-life functioning showed larger grey matter volume (increases) in the basal ganglia. This again seems to be the first time such finding has been described. However, the pathophysiological meaning of this result needs to be further elucidated, as current large-scale literature on structural alterations of the basal ganglia in SSD is rather inconclusive (6–8)

Furthermore, a higher positive symptom severity at baseline elicited worse post-intervention somatic health. This may result from the fact that patients with more severe positive symptoms may have received higher doses of antipsychotic medication that are known to have substantial somatic side effects such as weight gain (9).

Lastly, we obtained two counterintuitive results, indicating that a larger grey matter volume in the insula at baseline elicits higher positive symptom severity at post-intervention and suggesting that improvement in somatic health from pre- to post-intervention leads to lower daily-life functioning after the intervention. The first finding contradicts current large-scale evidence, as the insula – as part of the salience network – is known to be affected by decreases in grey matter volume linked to more severe psychopathology in psychiatric conditions (10,11). Given the exploratory and preliminary character of the current study, this finding needs to further evaluated and should not be interpreted at this point. The inverse association between worsening in somatic health and better daily-life functioning also contradicts current literature (12), but may be explainable by medication change during study participation. There may have been changes in antipsychotic treatment for some patients, leading to a worsening of somatic health (e.g. weight gain) accompanied by symptom improvements that also affect the GAF scale. As data on medication change was not available for all patients, we could not confirm this assumption.
